# Implementation of hospital-wide referral management by triage of non-urgent primary care referrals and experiences of specialists and general practitioners: a mixed methods study

**DOI:** 10.1101/2025.08.21.25334141

**Authors:** R.M.C. Pepping, R.C. Vos, M.C.G. Huijden, M. Crasborn, M.E. Numans, M.O. van Aken

## Abstract

**Introduction:** In the Netherlands, referral rates from primary to secondary care are rising due to an ageing population and complex healthcare needs, a challenge compounded by an on-going decline in the number of trained healthcare professionals. In this context, triage has shown promise in optimizing secondary care consultations. This hospital-wide study aimed to assess to what extent triaging non-emergent primary care referrals prevents outpatient consultations, as well as experiences of triage implementation by medical specialists and general practitioners (GP).

**Methods:** A mixed-methods study was conducted using routine care data from electronic health records (EHR) and semi-structured interviews. Referrals to 15 departments between August 2019 and July 2021 were included, with a six-month follow-up period. Referrals were assessed regarding the expected added value of secondary outpatient consultation and correctly chosen specialty. To gain insight into professionals’ experiences, interviews were conducted with GPs and with medical specialists from each participating department.

**Results:** A total of 109,953 primary care referrals were registered by participating departments. Of these, 4.262 (3.9%) were directed back to primary care, with redirection varying across departments (0% to 17.1%). Of the redirected referrals with six-months of follow-up, 274 of 3461 patients (7.9%) were re-referred for the same care need within this period. Qualitative findings showed overall positive experiences among medical specialists, with *major time investment* as the most important barrier to triage. GPs expressed more mixed feelings, with reported barriers including a sense of undermining of autonomy and lack of collaboration, although guidance and advice from specialists was much appreciated.

**Conclusion:** This study showed that a hospital-wide triage strategy can be effective in reducing outpatient consultations, with redirected referrals supported by advice and/or treatment guidance for the referring GP. Qualitative insights suggested that safeguarding mutual respect and cooperation between specialists and GPs needs to be addressed during the implementation of a triage system.

## Introduction

In the Netherlands referral rates from primary to secondary care have risen from 285 per 1000 patients in 2016 to 316 per 1000 patients in 2022 (1). Beginning in primary care, this growing demand is driven by an ageing and increasingly medically complex population, combined with a declining number of (available) healthcare professionals. These challenges place significant pressure on the healthcare system and underscore the need for better regulation of patient flows (2). General practitioners (GPs) in the Netherlands play a gatekeeper role in determining the secondary care needs of their patients. This formal and well-entrenched role within Dutch society is critical in managing healthcare demand and regulating referrals. However, GP referral rates vary considerably and reflect the diverse reasons underlying patient referral (3). Regulating demand via referral management is a difficult but important process in healthcare, which ideally should result in appropriate care in an appropriate setting.

Three categories of referral management interventions have previously been proposed: ‘education,’ ‘system/process change’, and ‘financial change’ (4, 5). A promising, proactive approach to referral management is triage, which falls under the second category ‘system/process change’. Triage is one proposed intervention to help address the referral process in a proactive manner and to safeguard the quality, affordability and accessibility of healthcare systems (6-8). A number of studies have demonstrated the effectiveness of triage in terms of patient safety and timely management, as well as patient and GP satisfaction (9-13). Potential for optimization and referral rate reduction was shown by two recent studies indicating that 10% to 28% of all incoming referrals can be redirected and handled with advice only, thus avoiding an in-person appointment (9, 11). Another study showed that 27% of incoming orthopaedic referrals were more appropriately handled by the rheumatology department (14). These data suggest that triage can improve appropriateness of referrals and help allocate referrals to the correct specialism. A weakness of the discussed studies was a focus on a specific complaint, condition or inclusion of a single medical specialty, rather than evaluation within a hospital-wide setting. Despite the promising results of triage reported to date, some care providers encounter (perceived) barriers to implementation and stress the importance of context-specific factors (12, 13).

In our hospital-wide study, referrals were assessed by medical specialists of outpatient departments, irrespective of the reason for referral, who then decided whether the referred patients required a hospital consultation. This study aimed to assess the extent to which triaging non-emergent referrals prevents outpatient consultations, as well as experiences of implementation among medical specialists and GPs.

## Methods

### Study design, setting and participants

A mixed-methods study, using routine care data from electronic health records (EHR) and semi-structured interviews. The hospital is located in an urban area in the Netherlands, serving a population of over 500,000 patients. All patients, both children and adults, with non-acute health complaints who were referred for an outpatient non-emergent consultation between August 2019 and July 2021 were included in the study. Referrals to the radiology, radiotherapy, and anaesthesiology departments were excluded, as were other outpatient referrals for GP diagnostic tests, since these were not assessed. Additionally, referrals for procedures outside the GPs’ scope were deemed ineligible for redirection. Interviews were conducted with medical specialists from each participating department, as well as GPs based in our region. Purposive sampling and personal networks were used to identify participants, with consideration given to their levels of involvement and experience with active triage.

### New assessment of referrals

In all departments, multiple dedicated specialists incorporated triaging into their work routine or allocated time in their schedule. All incoming referrals to their department were assessed, especially regarding the expected added value of the secondary care outpatient consultation and the correctness of the chosen specialty. As part of the procedure, patients triaged as requiring outpatient consultation were directed to the appropriate medical department and/or specialist. Redirection within the hospital occurred when consultation at another department was deemed more appropriate. When a referral to secondary care was considered unnecessary, a letter was sent and the medical specialist contacted the referring GP by telephone within 72 hours to explain the reasoning behind the decision to redirect and to offer advice (see Figure 1). In addition, the department of cardiology had the option to refer less complex patients expected to have minor or no cardiac problems to a local heart clinic for diagnosis and/or treatment.

**Figure 1.**
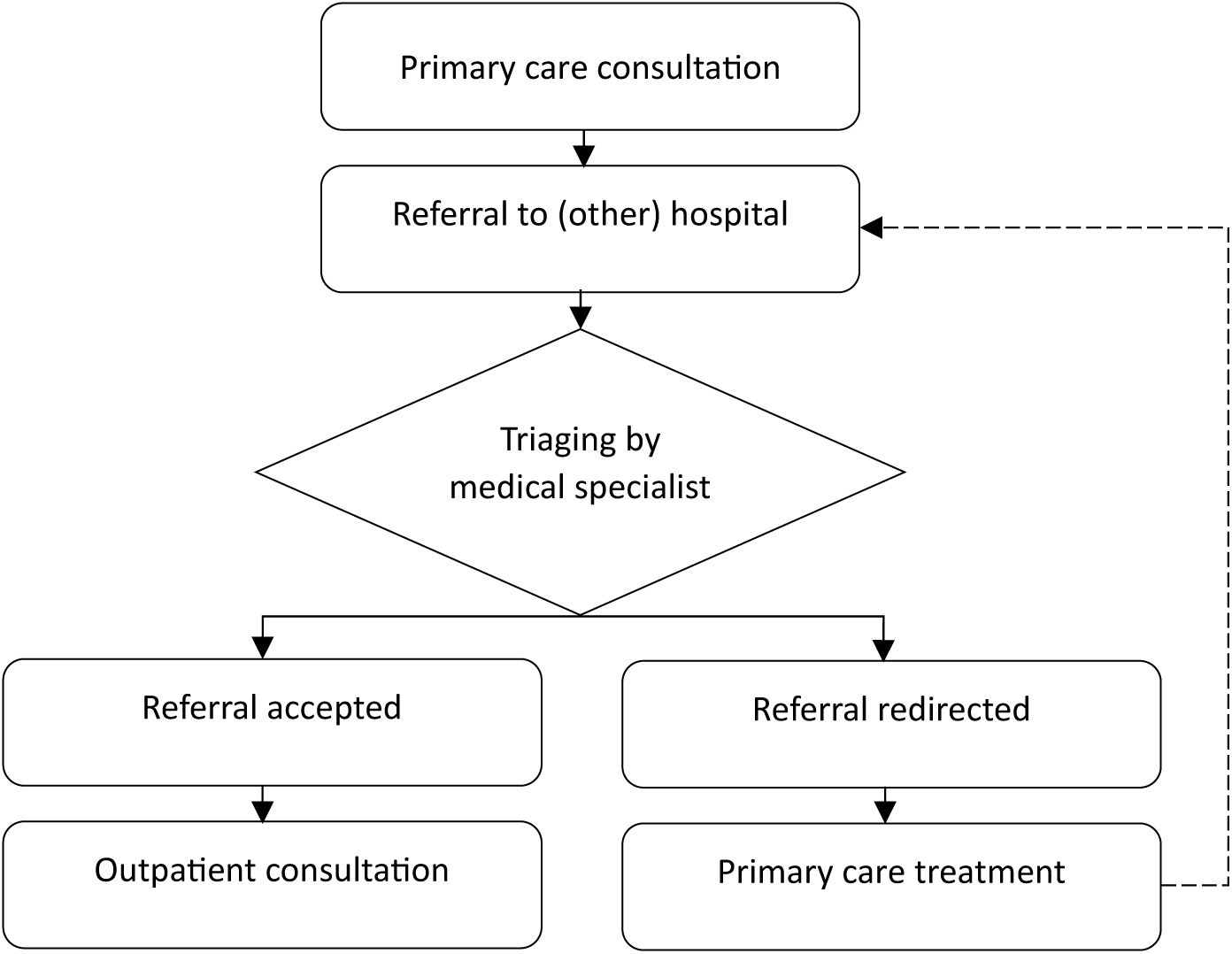
New triage process. Dotted line indicates a re-referral.

### Data collection

Quantitative data from the EHR were extracted from the hospital data warehouse. Every EHR with a recorded redirection between August 2019 and July 2020 was accessed in order to collect additional information on patient characteristics, referrer, urgency of the referral (regular, priority or urgent), medical specialty, medical complaint or condition, the reason for redirection if applicable, and whether or not an additional referral for the same complaint was made within the six-month follow-up period. After the first year, an interim analysis was conducted. Decreasing rates of adherence to the triage strategy were observed in most departments, with the exception of the cardiology and internal medicine departments. As a result, during the second year (from August 2020 to July 2021) additional data was collected solely from the cardiology and internal medicine departments (a period that coincided with the COVID-19 pandemic). Randomly chosen 100 EHRs were reviewed by two assessors to validate the decision regarding redirection. Categories of ‘reasons for redirection’ were based on the most common reasons found in literature and informed by the experience of a previous pilot triaging medical specialist, together resulting in 13 categories (Additional file 1).

#### Semi-structured interviews

To evaluate the experiences of stakeholders regarding implementation of the new triage strategy, semi-structured interviews were conducted using a topic list based on the Consolidated Framework for Implementation Research (CFIR) (15, 16). The CFIR encompasses a variety of domains designed to assess the implementation of new services, including (1) Intervention characteristics, (2) Outer setting, (3) Inner setting, (4) Characteristics of individuals, and (5) Process of implementation. This topic list was developed for this study (Supplementary file 1). Each department was contacted via email to inquire about their willingness to participate in an interview and to identify the medical specialist most closely involved with the triage process. This approach ensured that the specialist with the highest level of involvement in triage was interviewed. GPs in the region were predominantly approached through purposive and snowball sampling. The interviews were conducted by two (RP, MC) researchers between January 2022 and November 2022.

### Data analysis

The collected EHR data were analysed for frequencies and descriptive statistics using SPSS version 28.0.1.0. Using triage-based redirection rates, departments were categorised into three levels: high, medium, or low. The interviews were transcribed verbatim and analysed using ATLAS.ti, version 22.2.5.0. Thematic analysis was undertaken to identify recurring themes across the dataset, following the procedure outlined by Braun and Clarke (17). First, all transcripts were read and initial ideas noted to familiarise ourselves with the data. Next, initial codes were generated inductively to organise the data into meaningful groups. These codes were then sorted into potential themes and code groups. The data extracts within each theme were subsequently read and reviewed, with the potential themes refined by merging or breaking them down where necessary. Finally, clear definitions and names were assigned to the final themes.

To minimise the risk of bias, a second researcher was involved during this phase to independently code 10% of the interviews using the developed codebook. The coded interviews were then compared and discussed until consensus was reached regarding the themes and their definitions. We used the qualitative data to elucidate and explain the quantitative findings through an explanatory design (18-20).

## Results

### Referral rates

During this study a total of 109,953 primary care referrals were registered across the participating departments (Table 1). Of these incoming referrals, 4.262 (3.9%) were redirected back to primary care in accordance with the implemented strategy (Table 2). Departments with a high redirection rate, defined as over 5% of referrals redirected, included internal medicine, cardiology, rheumatology, and ophthalmology. Departments with moderate redirection rates, of between 1% and 5%, included dermatology, paediatrics, neurology, gastroenterology, pulmonology, and orthopaedics. Departments showing low redirection rates, with fewer than 1% of referrals redirected, included gynaecology, haematology, surgery, urology, and otorhinolaryngology (Table 2).

**Table 1.**
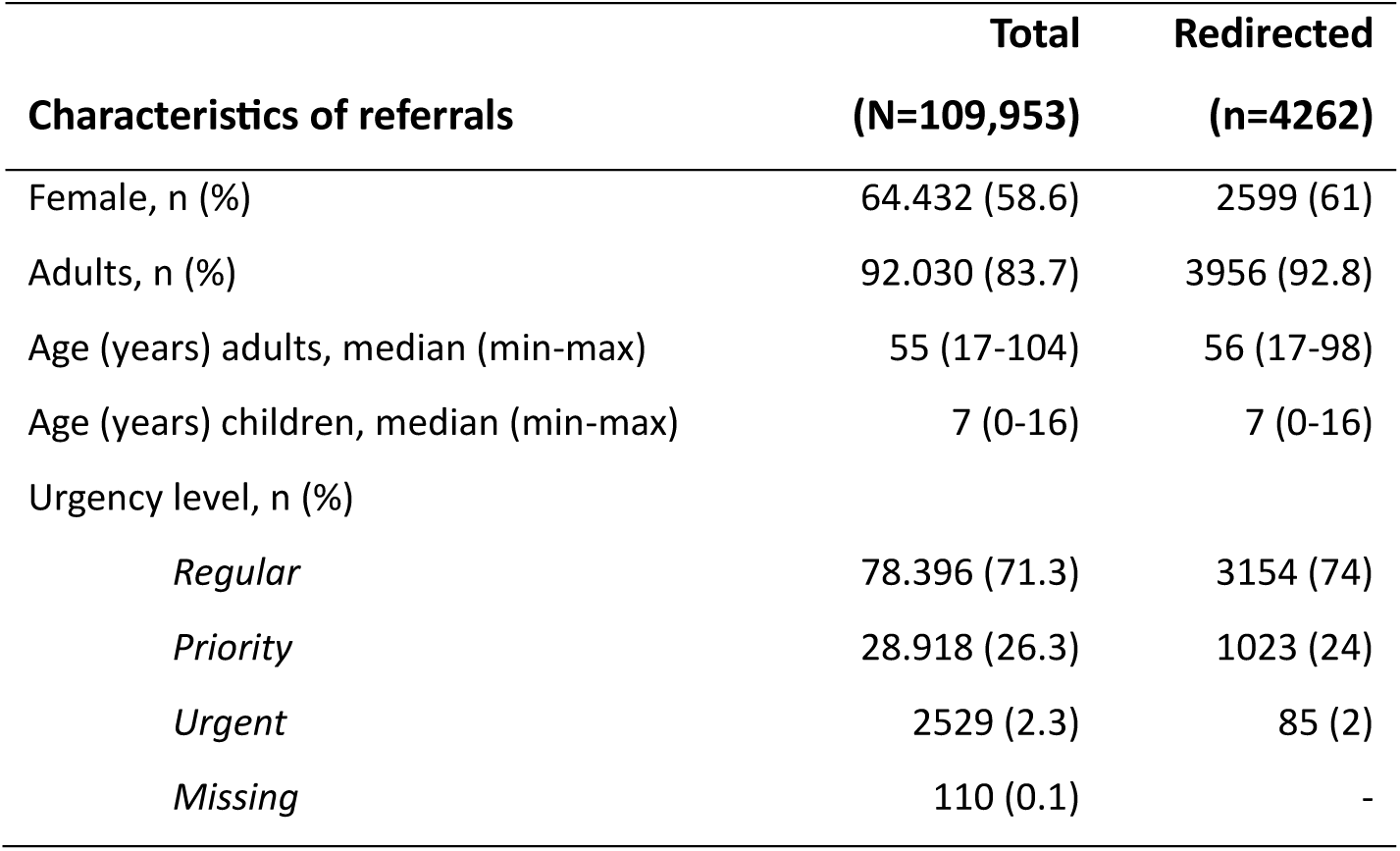
Characteristics of referrals. Urgency level was indicated by general practitioner.

| **Characteristics of referrals** | **Total**<br>**(N=109,953)** | **Redirected**<br>**(n=4262)** |
| --- | --- | --- |
| Female, n (%) | 64.432 (58.6) | 2599 (61) |
| Adults, n (%) | 92.030 (83.7) | 3956 (92.8) |
| Age (years) adults, median (min-max) | 55 (17-104) | 56 (17-98) |
| Age (years) children, median (min-max) | 7 (0-16) | 7 (0-16) |
| Urgency level, n (%) |  |  |
| *Regular* | 78.396 (71.3) | 3154 (74) |
| *Priority* | 28.918 (26.3) | 1023 (24) |
| *Urgent* | 2529 (2.3) | 85 (2) |
| *Missing* | 110 (0.1) | - |

**Table 2.** Assessment of effectiveness of active triage per medical department.

| **Department, N (%)** | **Total**<br>**(N=109,953)** | **Redirected**<br>**(n=4262)** |
| --- | --- | --- |
| Internal Medicine* | 5835 | 998 (17.1) |
| Cardiology* | 7446 | 1.068 (14.3) |
| Rheumatology | 2989 | 215 (7.2) |
| Ophthalmology | 11.883 | 664 (5.6) |
| Dermatology | 9617 | 387 (4.0) |
| Paediatrics | 6356 | 213 (3.4) |
| Neurology | 12.040 | 332 (2.8) |
| Gastroenterology | 4267 | 109 (2.6) |
| Pulmonology | 2257 | 54 (2.4) |
| Orthopaedics | 7438 | 96 (1.3) |
| Gynaecology | 9350 | 71 (0.8) |
| Haematology | 503 | 4 (0.8) |
| Surgery | 8078 | 25 (0.3) |
| Urology | 6838 | 23 (0.3) |
| Otorhinolaryngology | 14.696 | 3 (0.0) |

*\*Two-year follow-up*

### Reason to redirect

Reasons for redirection varied across departments, with *Limited contribution of secondary care* most frequently reported in 7 out of 15 departments (14.1%), followed by *Additional advice to the GP* (10.5%) and *Diagnosis already known/continue treatment at a known department or hospital* (7.6%) (Table 3). For the cardiology department, *Redirection to the heart clinic*, was the most prevalent reason for redirection (Table 3 and Additional file 1 (Table 5)). Rheumatology and paediatrics showed the highest rates of internal redirection (Additional file 1 (Table 5)), with paediatrics primarily redirecting cases internally due to in-house agreements to concentrate allergy care within the dermatology department. However, following interim analyses at year one, in 801 cases the reason for redirection was omitted, mostly in departments with already moderate to low redirection rates.

**Table 3.** Reasons for redirection, hospital wide. GP: general practitioner.

| Reason to redirect, N (%) | Total<br>(N=3461) |
| --- | --- |
| Limited contribution of secondary care | 603 (14.1) |
| Additional advice to GP | 446 (10.5) |
| Diagnosis already known | 323 (7.6) |
| Redirection within the hospital | 251 (5.9) |
| Laboratory error, laboratory results within normal range | 158 (3.7) |
| Additional examination by GP | 144 (3.4) |
| Primary care guidelines not followed | 149 (3.5) |
| Referral cancelled by patient | 65 (1.5) |
| Referral to private practice | 38 (0.9) |
| Advice via teleconsultation | 94 (2.2) |
| Referral to heart clinic | 829 (19.5) |
| Other | 248 (5.8) |
| Unknown | 113 (2.7) |

**Table 4.** Re-referrals per department.

| Department, N (%) | Re-referred<br>(n=274) | Total redirected per<br>department (N=3461) |
| --- | --- | --- |
| Internal medicine* | 86 (8.6) | 998 |
| Cardiology* | 48 (4.5) | 1068 |
| Rheumatology | 11 (8.5) | 129 |
| Ophthalmology | 45 (11.2) | 403 |
| Dermatology | 14 (6.5) | 214 |
| Paediatrics | 13 (7.6) | 171 |
| Neurology | 29 (14.7) | 197 |
| Gastroenterology | 7 (12.1) | 58 |
| Pulmonology | 5 (15.2) | 33 |
| Orthopaedics | 7 (7.2) | 96 |
| Gynaecology | 5 (8.2) | 61 |
| Haematology | 0 (0.0) | 3 |
| Surgery | 2 (15.4) | 13 |
| Urology | 1 (6.7) | 15 |
| Otorhinolaryngology | 1 (50.0) | 2 |

\* Two-year follow-up

**Table 5.**
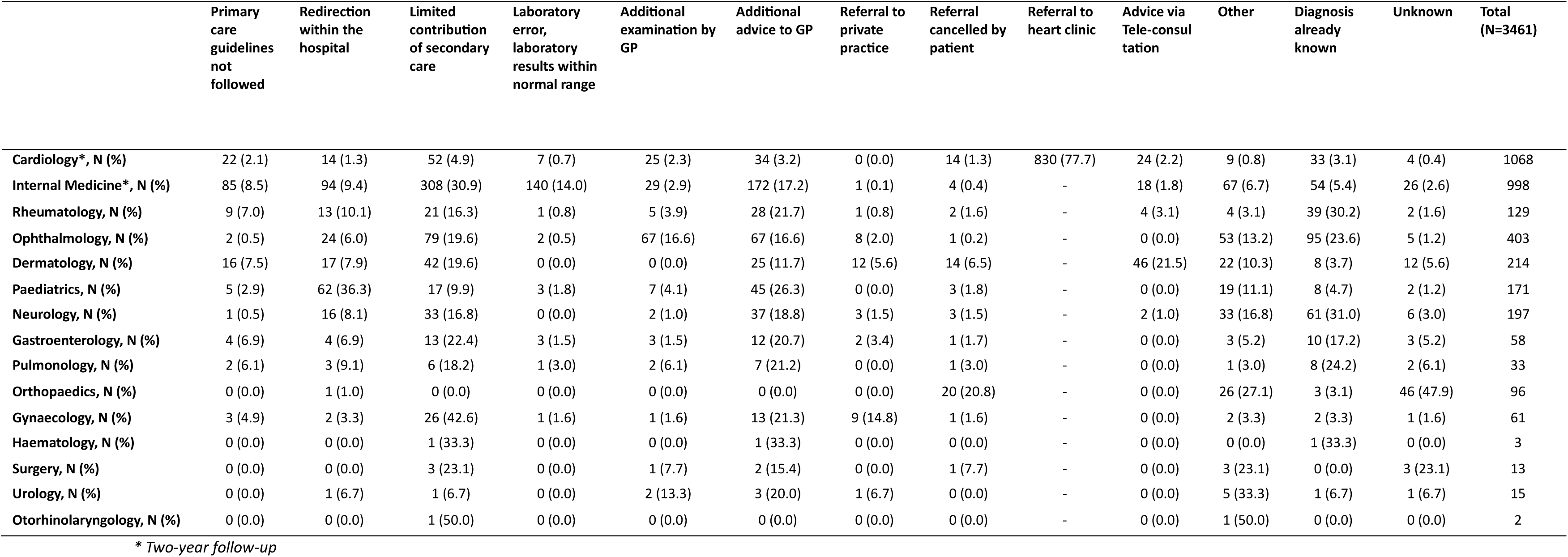
Department specific reasons to redirect. GP: general practitioner.

|  | Primary care guidelines not followed | Redirection within the hospital | Limited contribution of secondary care | Laboratory error, laboratory results within normal range | Additional examination by GP | Additional advice to GP | Referral to private practice | Referral cancelled by patient | Referral to heart clinic | Advice via Tele-consultation | Other | Diagnosis already known | Unknown | Total (N=3461) |
| --- | --- | --- | --- | --- | --- | --- | --- | --- | --- | --- | --- | --- | --- | --- |
| Cardiology*, N (%) | 22 (2.1) | 14 (1.3) | 52 (4.9) | 7 (0.7) | 25 (2.3) | 34 (3.2) | 0 (0.0) | 14 (1.3) | 830 (77.7) | 24 (2.2) | 9 (0.8) | 33 (3.1) | 4 (0.4) | 1068 |
| Internal Medicine*, N (%) | 85 (8.5) | 94 (9.4) | 308 (30.9) | 140 (14.0) | 29 (2.9) | 172 (17.2) | 1 (0.1) | 4 (0.4) | - | 18 (1.8) | 67 (6.7) | 54 (5.4) | 26 (2.6) | 998 |
| Rheumatology, N (%) | 9 (7.0) | 13 (10.1) | 21 (16.3) | 1 (0.8) | 5 (3.9) | 28 (21.7) | 1 (0.8) | 2 (1.6) | - | 4 (3.1) | 4 (3.1) | 39 (30.2) | 2 (1.6) | 129 |
| Ophthalmology, N (%) | 2 (0.5) | 24 (6.0) | 79 (19.6) | 2 (0.5) | 67 (16.6) | 67 (16.6) | 8 (2.0) | 1 (0.2) | - | 0 (0.0) | 53 (13.2) | 95 (23.6) | 5 (1.2) | 403 |
| Dermatology, N (%) | 16 (7.5) | 17 (7.9) | 42 (19.6) | 0 (0.0) | 0 (0.0) | 25 (11.7) | 12 (5.6) | 14 (6.5) | - | 46 (21.5) | 22 (10.3) | 8 (3.7) | 12 (5.6) | 214 |
| Paediatrics, N (%) | 5 (2.9) | 62 (36.3) | 17 (9.9) | 3 (1.8) | 7 (4.1) | 45 (26.3) | 0 (0.0) | 3 (1.8) | - | 0 (0.0) | 19 (11.1) | 8 (4.7) | 2 (1.2) | 171 |
| Neurology, N (%) | 1 (0.5) | 16 (8.1) | 33 (16.8) | 0 (0.0) | 2 (1.0) | 37 (18.8) | 3 (1.5) | 3 (1.5) | - | 2 (1.0) | 33 (16.8) | 61 (31.0) | 6 (3.0) | 197 |
| Gastroenterology, N (%) | 4 (6.9) | 4 (6.9) | 13 (22.4) | 3 (1.5) | 3 (1.5) | 12 (20.7) | 2 (3.4) | 1 (1.7) | - | 0 (0.0) | 3 (5.2) | 10 (17.2) | 3 (5.2) | 58 |
| Pulmonology, N (%) | 2 (6.1) | 3 (9.1) | 6 (18.2) | 1 (3.0) | 2 (6.1) | 7 (21.2) | 0 (0.0) | 1 (3.0) | - | 0 (0.0) | 1 (3.0) | 8 (24.2) | 2 (6.1) | 33 |
| Orthopaedics, N (%) | 0 (0.0) | 1 (1.0) | 0 (0.0) | 0 (0.0) | 0 (0.0) | 0 (0.0) | 0 (0.0) | 20 (20.8) | - | 0 (0.0) | 26 (27.1) | 3 (3.1) | 46 (47.9) | 96 |
| Gynaecology, N (%) | 3 (4.9) | 2 (3.3) | 26 (42.6) | 1 (1.6) | 1 (1.6) | 13 (21.3) | 9 (14.8) | 1 (1.6) | - | 0 (0.0) | 2 (3.3) | 2 (3.3) | 1 (1.6) | 61 |
| Haematology, N (%) | 0 (0.0) | 0 (0.0) | 1 (33.3) | 0 (0.0) | 0 (0.0) | 1 (33.3) | 0 (0.0) | 0 (0.0) | - | 0 (0.0) | 0 (0.0) | 1 (33.3) | 0 (0.0) | 3 |
| Surgery, N (%) | 0 (0.0) | 0 (0.0) | 3 (23.1) | 0 (0.0) | 1 (7.7) | 2 (15.4) | 0 (0.0) | 1 (7.7) | - | 0 (0.0) | 3 (23.1) | 0 (0.0) | 3 (23.1) | 13 |
| Urology, N (%) | 0 (0.0) | 1 (6.7) | 1 (6.7) | 0 (0.0) | 2 (13.3) | 3 (20.0) | 1 (6.7) | 0 (0.0) | - | 0 (0.0) | 5 (33.3) | 1 (6.7) | 1 (6.7) | 15 |
| Otorhinolaryngology, N (%) | 0 (0.0) | 0 (0.0) | 1 (50.0) | 0 (0.0) | 0 (0.0) | 0 (0.0) | 0 (0.0) | 0 (0.0) | - | 0 (0.0) | 1 (50.0) | 0 (0.0) | 0 (0.0) | 2 |

\* Two-year follow-up

### Re-referrals

Of the 3.461 redirected referrals with data available on six-month follow-up, 274 (7.9%) patients had a re-referral for the same care need within six months after redirection of the first referral (see Table 4). Most re-referrals (n=221, 80.6%) were addressed to the same medical department as the initial referral, and almost 30% were re-referred within 30 days of the original referral.

### Interviews

To understand contextual factors that influenced triage strategy implementation, qualitative data were used in an explanatory sequential design. (18) In total, 15 interviews were conducted with medical specialists and 13 interviews with GPs. Medical specialists had a mean age of 48.1 years, with on average 15.4 years of work experience, while GPs had a mean age of 44.4 years, with 14.5 years of GP work experience.

We identified 17 unique barriers and 15 unique facilitators based on the CFIR framework. Some determinants were valued differently between departments with high and moderate to low redirection rates. Furthermore, medical specialists and GPs sometimes differed concerning the same determinants (see appendix 1 and 2 for the code tree and code book resulting from the thematic analysis). The principal barriers and facilitators are further discussed below, together with the matching determinant or overarching domain.

#### General and mixed determinants

As envisioned within the strategy, triage was performed mainly by medical specialists, although three departments mentioned shifting responsibility to residents and medical secretaries [adaptability]. The importance of triage by a medical specialist was stressed mainly by departments with high redirection rates, whereas other departments had mixed opinions. Some felt that when responsibility was shifted away from medica specialists, triage should be strictly protocolized, potentially resulting in restricted opportunities for triage. Departments also varied in how triage was incorporated into daily routines [compatibility], with some scheduling a dedicated time of day, while others triaged all referrals at the end of their shift. Multiple specialists indicated that triage felt like another task added to an already busy schedule (Quote A). On the other hand, departments with low redirection rates adhered less closely to the strategy and more often mentioned shifting triaged referrals to (electronic) teleconsultations or other hospital departments, possibly explaining the lower rates of redirection to primary care [intervention domain].

Both medical specialists and GPs stressed that clear communication was the most important factor in the success of the strategy. Medical specialists prefer GPs to provide a detailed explanation of the referral, but opinions varied on whether patient preferences should be included, as some believed this could negatively influence triage decisions and potentially affect quality of care [executing] (Quote B). In addition, some medical specialists advocated use of referral templates for specific care needs to ensure inclusion of essential information. However, GPs expressed concerns that templates might diminish the nuances of referrals and act as a barrier to making referrals [outer setting domain].

> *(A) “… it is getting labour-intensive, which makes us sometimes think: just send the patient to the hospital.” – Medical specialist*
>
> *(B) “…in the end I decided to re-refer her to the internist, who can take a broader view.” – GP*

#### Facilitators

An important facilitator of triage implementation by medical specialists was the option to redirect referrals to primary care [Intervention domain]. In addition, assigning the most qualified specialist, indicating the preferred waiting time for consultation, and initiating diagnostic tests (i.e., laboratory and radiology) prior to consultation were identified as important facilitators of triage by some departments (Quote C). Another facilitator for departments with the most redirections was the positive perception of a shift in the outpatient case mix, with more patients presenting with complex health issues. Specialists expressed enthusiasm regarding this shift, remarking that treating a complex population was why they had become specialists. Interestingly, these departments felt they were controlling their workload by attracting only patients with complex health issues, whereas departments with moderate and low redirections expressed the view that less complicated health complaints helped to ease their workload [Attitude] (Quote D).

The primary facilitators for GPs were the guidance and advice provided by medical specialists in the accompanying redirection letters [Intervention domain]. This support enabled GPs to better explain issues to their patients and offer appropriate guidance (Quote E). The preferred method of communication was a phone call together with a redirection letter, as in some cases the letter alone was perceived as a rebuff [Outer setting domain] (Quote F). Nonetheless, after a phone call most GPs agreed to the redirection, and an additional facilitator was the positive learning experience following explanation of the redirection.

> *(C) “Look, triaging is partly rejection, say five percent, but the rest is more triaging in terms of am I already doing diagnostic tests and in what time period.” – Medical specialist*
>
> *(D) “And it also makes our profession more enjoyable, because yes, I enjoy treating more complex pathologies. And now I have more time for those complex patients.” – Medical specialist*
>
> *(E) “… it’s not that they need to see the cardiologist face-to-face. It’s more about the cardiologists’ expertise. Then they’re satisfied.” – GP*
>
> *(F) “I would rather have received an explanatory phone call, because then I could ask why this is the case.” – GP*

#### Barriers

In the opinion of the medical specialists, they faced three crucial barriers to performing triage as intended within the developed strategy. The first barrier was the major time investment required to conduct triage, which negatively influenced its outcomes [Implementation climate]. In terms of efficient use of time, the constant switching between communication channels, i.e., redirection letters versus phone calls led to mixed feelings, as for example one department decided to only call rather than write redirection letters to referring GPs due to greater perceived efficiency. The misalignment between GP’s and specialist’s work schedules was also frequently mentioned as further hampering efficient triage. This time investment was also cited as a reason to discontinue triage, especially in departments with low redirection rates (see Table 2). One department explicitly stated that performing triage consumed the specialist’s valuable time which could be better spent (Quote G). The second barrier was the quality and content of primary care referral letters [Executing]. These were considered essential by all medical specialists, yet necessary key information – such as relevant medical details, the (anticipated) diagnosis, and a clear question – was often lacking, again hampering effective triage (Quote H). A third barrier, highlighted by nearly all departments, was related to the already substantial workload of GPs and the perception that patients were being "delegated" or "dumped" on them [Characteristics domain]. Factors such as the (potential) lack of appropriate follow-up options or limited GP expertise were perceived as beyond the specialist’s control but still impacted triage outcomes. Additionally, two minor barriers were noted: the high volume of monthly incoming referrals and the perceived necessity of performing a physical examination regardless.

Some GPs felt that redirected referrals undermined their autonomy, as they thought specialists disregarded their perspective and focused solely on medical aspects [Attitude] (Quote I). Secondly, they also argued that this approach failed to foster teamwork and discouraged collaboration [Cosmopolitanism]. Furthermore, GPs highlighted the need for a second opinion before continuing care in a primary care setting, an issue they viewed as essential to maintaining care quality. Another barrier was the patient’s perception of quality of care once a GP had indicated a lack of expertise but the referral was rejected by the hospital [Outer setting] (Quote J).

> *(G) “No, I prefer to see all patients. I would rather not reject anyone, but it’s kind of imposed on us.” – Medical specialist*
>
> *(H) “Yes, a more detailed referral allows better targeting of diagnostic tests and more efficient use of our resources. With a broad referral that is much more difficult.” – Medical specialist*
>
> *(I) “… because I refer a patient for a reason. That also means I feel I don’t have the expertise myself. But these days you almost have to beg a specialist to see your patient.” – GP*
>
> *(J) “Yes, hearing ‘we can’t treat this patient’ sometimes feels like a slap in the face. Then I think: I am still stuck with the patient and what am I supposed to do now?” – GP*

## Discussion

This study aimed to assess the extent to which triaging non-emergent primary care referrals avoids outpatient consultations, as well as assess the experiences of implementation among medical specialists and GPs. This referral strategy was conceived as an attempt to improve the system and regulate growing demand. Our results showed a hospital-wide reduction of almost 4% and department-specific reductions of up to 17.1% in outpatient consultations. These referrals were solved in a different way by triage and guidance of GPs in their decisions. A re-referral rate of only 7.9% within a six-month follow-up period indicated that this is a safe, satisfactory strategy in the vast majority of cases. Based on our interviews, adequate triage was significantly impacted by the quality of information in the referral letter. This finding is in agreement with other reports, suggesting that referral letters are often inconsistent and lack essential information (14, 21, 22). Several departments, particularly those showing moderate to low redirection, indicated that the main benefit of triage was the early initiation of diagnostic tests, rather than substitution of avoidable hospital care by primary care.

As regards the effectiveness of triage by medical specialists in preventing outpatient consultations, our findings partially align with previous research. Earlier studies of referrals redirected to primary care reported percentages of up to 71%; however, these results were often confined to a single medical specialty (7, 9-11, 14). Our goal was not only to reduce consultations, but also to provide advice and feedback to GPs, offer reassurance concerning ongoing treatments, and ensure that patients were seen by the appropriate specialist during their initial visit.

We found considerable variation in percentages of redirected referrals across and between departments, a finding similar to other reports where 10% to 45% and 20-35% of referrals were considered by secondary care specialists to have limited added value (23, 24). This “limited contribution of secondary care” was also the main reason for re-directing referrals in our study. Although this assessment represents the initial opinion of the medical specialist, most GPs agreed with this assessment after a phone call and a formal letter including treatment advice.

Nevertheless, not all departments felt it necessary to call GPs and provide treatment guidance in a formal letter after declining a referral. This was seen in some specialities as unnecessary and inefficient, an opinion particularly evident in surgical specialties. This may have led to under-reporting of true redirection rates, as our statistics (3.9%) were based on redirection letters extracted from the EHR, possibly suggesting that the real number of redirected referrals in some departments was higher than reported here. Between-department differences in fidelity to the triage strategy might also explain the difference in perspectives between medical specialists and GPs. While specialists reported active involvement, GPs perceived the process as top-down and asymmetrical, and characterised by limited two-way communication. GPs perceived this lack of communication as having a negative impact on collaboration, personal and professional relationships. These findings align with previous qualitative research on referral processes: for example, in-depth interviews with Canadian GPs revealed feelings of being overwhelmed by workloads imposed by medical specialists (25, 26). Conversely, a study confined to the paediatric specialty indicated that GPs were very satisfied with adjustments to referrals, with an observed educational benefit (10).

The perceived efficiency of triage by specialists may partly account for variations between departments. Differences in factors such as the monthly volume of incoming referrals and the reliance on physical examinations influence the feasibility and outcomes of triage. These variations have also sparked ongoing debates concerning who is best suited to handle triage. One study demonstrated that experienced neurology nurses can effectively “flag” referrals for subsequent evaluation by neurologists (7). This approach could be one solution, as multiple medical specialists reported that triage consumes too much of their time while yielding minimal or no change in their case mix. In reality, the same study indicated that a 20-minute time investment in triage could free up one hour in the outpatient clinic schedule, allowing specialists to focus on patients requiring their expertise.

In our study, we observed a re-referral rate of only 7.9% over the six-month follow-up period, which compares favourably with the 15% and 30% rates reported in two studies, both with a one-year follow-up (7, 9). This may suggest that both GPs and patients benefit from the advice provided. However, it remains unclear whether “our” patients were subsequently referred to other hospitals within the region. It is worth mentioning that among the GPs interviewed, there was a general tendency not to refer patients to other hospitals. We also found that most re-referrals occurred within 30 days of the initial referral, which may indicate that when a GP or patient strongly favours a referral, it tends to happen quickly and to the same hospital. Unfortunately, no data were available regarding clinical decisions in these cases, i.e. did patients get a hospital appointment or were they redirected once again. During interviews with both GPs and medical specialists it appeared that the majority of these re-referred patients eventually received a scheduled appointment. This conclusion is supported by a small Dutch study of nephrology referrals, which reported that only six of 96 redirected patients were re-referred (27). All six received an appointment, but four were seen only once at the hospital before being referred back to their GP once again. This implies that secondary care was unnecessary in these cases, but our experience from both viewpoints suggests that a hospital appointment can sometimes support both the GP and the patient in continuing treatment within primary care.

## Strengths and limitations

A major strength of our study was the hospital-wide perspective covering all major outpatient departments and potential diagnoses, with results based on a very large dataset over a two-year period. Differences in referral and redirection rates were found among departments and enriched with qualitative insights, with perspectives on experiences and differences in implementation enhanced by this mixed methods approach. This study also improves our recognition and understanding of department-specific barriers, allowing context-specific adaptations to strategy, an interpretative component that was missing from earlier studies, which primarily focused on measuring effectiveness and/or safety. As a result of the decision to stop collecting data following the interim analysis, a limitation is that missing data in the second implementation year were not random. However, in our opinion similar findings are to be expected based on the one-year analyses. Another possible limitation was the lack of data from other regional hospitals, resulting in uncertainty regarding whether patients were subsequently referred to other health care providers. This underlines the importance of a regional approach. Further, as purposive and snowball sampling were used to include GPs in our study some selection bias may have been present.

## Conclusion

This study presents a practical, hospital-wide triage strategy for medical specialists wishing to regulate referrals. Our findings indicate a potential hospital-wide reduction of almost 4% and up to 17.1% in certain departmental outpatient consultations, highlighting opportunities to safely redirect patients to primary care, ensure consultations with the appropriate specialist, and initiate required examinations by GPs before consultation. Departments with high triage adoption rates valued the fact that hospital consultations better focused on patients requiring their expertise, while those with moderate or low triage adoption valued the early initiation of diagnostic tests (e.g., laboratory and radiology). Although GPs experienced more barriers than facilitators compared to medical specialists, they nevertheless expressed enthusiasm for the educational benefits and collaboration with specialists. The experiences of medical specialists and GPs described here provide valuable insights for clinicians and policymakers aiming to improve referral pathways.

## Declarations

### Ethical considerations

The study was approved by the Medical Ethics Committee of Leiden The Hague Delft (reference N21.093). The protocol was subsequently presented to the hospital ethics committee, who approved the protocol (reference T21-077). All interviewees gave written informed consent to participate in the study. All methods were performed in accordance with the Declaration of Helsinki.

### Consent for publication

Not applicable.

### Availability of data and materials

The datasets used and/or analysed during the current study are available from the corresponding author on reasonable request. Transcribed interviews are not available due to privacy laws.

### Competing interests

The authors declare that they have no competing interests.

### Funding

Not applicable

### Author contributions

Conception and design: RP, RV, MC, MvA. Acquisition of data: RP, MC. Analysis and interpretation of data: RP, MH. Drafting the article: RP, MH, RV, MvA. Final approval: RP, RV, MH, MC, MN, MvA.

## Acknowledgements

We would like to thank Mathilde Overtoom with her support during data extraction of the electronic health records. We also would like to express our sincere gratitude to all interviewed medical specialists and general practitioners for providing us with valuable insights and their (very) honest opinions.

## List of abbreviations

CFIR: Consolidated Framework for Implementation Research
EHR: Electronic health record
GP: General practitioner

## Appendix 1: Code tree

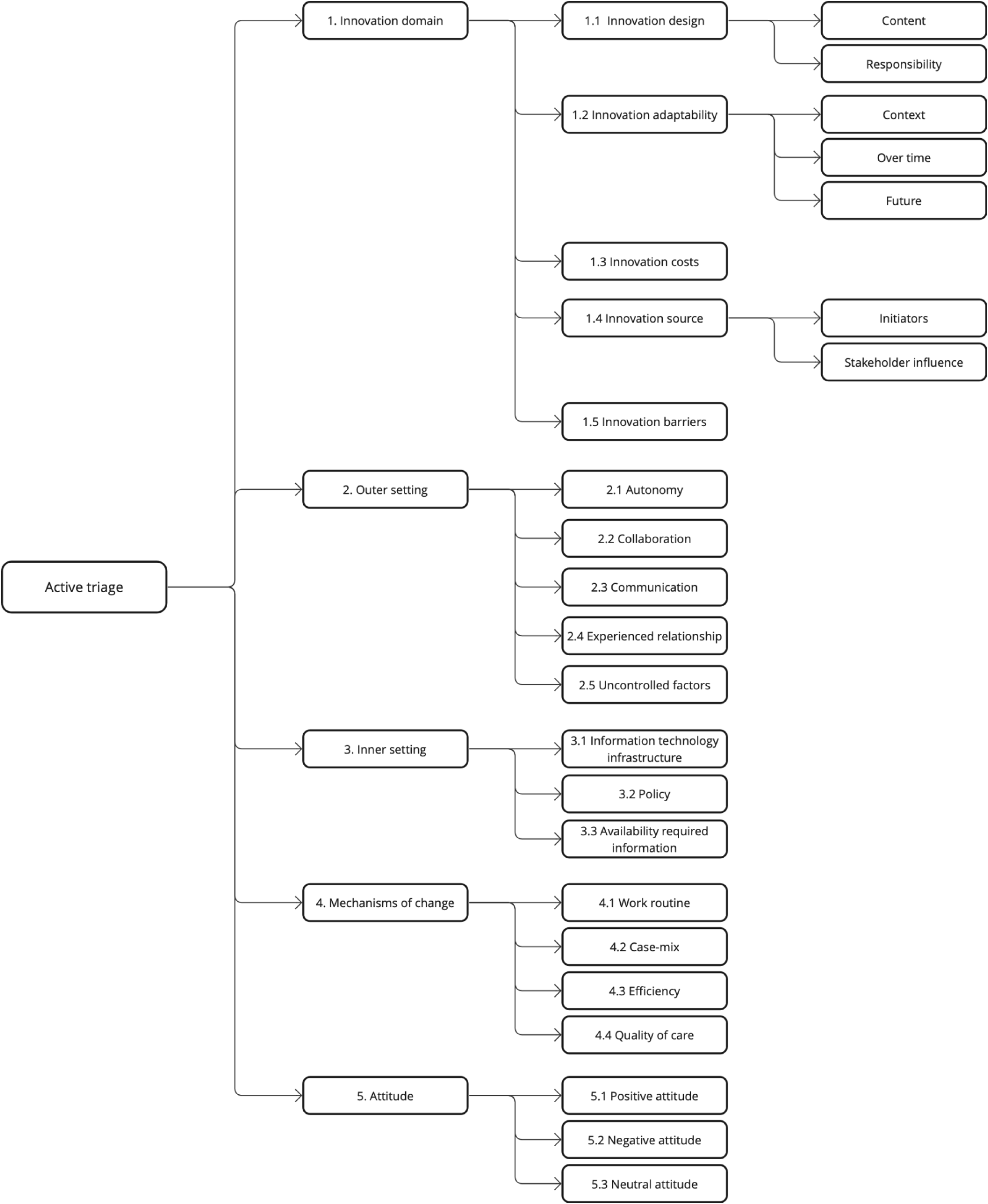

## Appendix 2: Code book

| Code group | Code name | Comment |
| --- | --- | --- |
| Innovation domain | 1.1 Innovation design: Content | The design of the intervention, including the various elements and how it is bundled. |
|  | 1.1 Innovation design: Responsibility | Delegation of tasks to execute the various elements of the intervention. |
|  | 1.2 Innovation adaptability: Context | How the intervention is modified, tailored, or refined to fit the local context/department and/or needs. |
|  | 1.2 Innovation adaptability: Over time | How the intervention is modified, tailored, or refined during the study period. |
|  | 1.2 Innovation adaptability: Future | How the intervention can be modified, tailored, or refined in the future to fit the local context/department and/or needs. |
|  | 1.3 Innovation costs | Financial, material, and personnel investments in the intervention. |
|  | 1.4 Innovation source: Initiators | The group that is perceived as initiating, developing, and/or sponsoring the use of the intervention. |
|  | 1.4 Innovation source: Stakeholder influence | The degree to which stakeholders (besides the initiators) are involved in the development and/or implementation of the intervention. |
|  | 1.5 Innovation barriers | Factors related to how the intervention is bundled and presented that are negatively influencing the practical execution of the intervention. |
| Outer setting | 2.1 Autonomy | The organization is networked with external entities: Formal and informal role/autonomy divisions between the organization and external entities. |
|  | 2.2 Collaboration | The organization is networked with external entities: Formal and informal collaboration structures between the organization and external entities. |
|  | 2.3 Communication | The organization is networked with external entities: Formal and informal information sharing practices between the organization and external entities. |
|  | 2.4 Experienced relationship | The organization is networked with external entities: Experienced influence of the intervention on the external relationship as expressed by involved stakeholders. |
|  | 2.5 Uncontrolled factors | Factors influencing the implementation, execution and/or outcome of the intervention that are outside the scope of the organization, including patient empowerment and practice variance. |
| Inner setting | 3.1 Information technology infrastructure | Technological systems for documentation, data storage, management, reporting, and communication support the implementation and/or execution of the intervention. |
|  | 3.2 Policy | Formal and informal information sharing practices concerning the intervention within the inner setting, including rules and related tangible and/or intangible incentives. |
|  | 3.3 Availability of required information | Needed information is available and accessible to execute the intervention. |
| Mechanisms of change | 4.1 Work routine | The impact of the intervention on the care delivery process, including changes in the daily work routine of stakeholders involved and/or affected by the intervention. |
|  | 4.2 Case-mix | The impact of the intervention on the characteristics of the outpatient population, including the role of a learning curve among GPs and how change is perceived by the involved stakeholders of the organization. |
|  | 4.3 Efficiency | The impact of the intervention on the (in)efficiency of the care delivery process. |
|  | 4.4 Quality of care | The impact of the intervention on the delivered quality of care. |
| Attitude | 5.1 Positive attitude<br>5.2 Negative attitude<br>5.3 Neutral attitude |  |

## Notes

### Competing Interest Statement

The authors have declared no competing interest.

### Funding Statement

This study did not receive any funding.

